# Suicide among older adults in Austria

**DOI:** 10.64898/2026.08.02.26359540

**Authors:** Anna Schultz, Emilise Pötz, Carlos Watzka, Christian Jagsch, Thomas Niederkrotenthaler, Erwin Stolz, Annette Erlangsen

## Abstract

**Background:** Suicide rates are highest among older adults. Yet, little is known regarding differences in suicide risk of older adults in Austria. This nationwide, retrospective cohort study aimed to characterize suicides among older adults in Austria during 2014-2023 and examine differences those who were young-old (65-74), middle-old (75-84), and oldest-old (85 +).

**Methods:** We applied a cohort design to individual-level linkage data on all residents in Austria aged 65 years and older during 2014-2022 (*N* = 2,442,939). We compared characteristics of individuals who died by suicide and other causes of death, as well as by age group (i.e., young-old, middle-old, oldest-old) using trend tests and odds ratios. We calculated crude incidence rates of suicide per age group and sex.

**Results:** During 2014-2023, 4,724 older adults died by suicide in Austria. The overall suicide rate was 23.4 per 100,000 person-years, while males had higher rates than females. Suicide rates increased with age and peaked among the oldest-old (33.2/100,000). Hanging, firearms, and jumping were the predominant methods, and the prevalence of hanging increased relative with increasing age. With increasing age, also widowhood and cardiovascular and genitourinary disorders were more prevalent among those who died by suicide.

**Conclusions:** In Austria, oldest-old adults have the highest suicide rates, especially for males. whereas rates were considerably lower and did not change throughout old age for females. Distinct differences in suicide rates with respect to marital status, health conditions, and suicide methods. This emphasizes the need for preventive strategies to target older adults at different stages of life.

## Background

Suicide is a relevant public health issue [1,2] and a major preventable cause of death worldwide, accounting for more than 700.000 deaths in 2021 [3]. Although overall suicide rates have declined over recent decades, rates among older adults remain alarmingly high, particularly among those aged 70 years and older [4]. This is also observed in Austria (Supplemental Figure 1); Austrian males aged 85 and above had the second-highest suicide rate of all age groups in the EU countries based on the most recent available data (114.8 per 100,000 person-years) [5]. Yet suicides in later life have received relatively little attention [6–9], and further research is needed to inform prevention strategies [9,10].

A number of risk factors have been linked to suicide in older adults, including higher age, male sex, low education [11], physical diseases [11–13], disabilities [14,15] mental disorders [11,16], and divorce or bereavement [11,17]. However, older adults do not constitute a homogeneous group, and the impact by potential risk factors might vary by age [18–20]. Koo and colleagues [18], for example, found that the role of psychiatric disorders, legal and financial difficulties, and interpersonal conflicts as well as suicide attempts diminished relative with increasing age, while physical health problems, bereavement, and exposure to stressful life events became increasingly prevalent in later life.

To guide prevention strategies, a clearer understanding of suicide-related characteristics in later life is needed. Accordingly, this nationwide retrospective cohort study aimed to analyze whether sociodemographic factors, comorbidities, and suicide methods varied between 1) older adults who died by suicide and those who died from other causes, and 2) young-old (65-74 years), middle-old (75-84 years) and oldest-old (85 years and older) individuals who died by suicide in Austria during 2014 and 2023

## Methods

### Design, setting and data sources

We included all individuals aged 65 years or older living in Austria between 2014 and 2022 who were followed up until December 31, 2023. Data was obtained via the Austrian Micro Data Center (AMDC) for the years 2014-2023, where covariates from the following individual-level registers were linked via pseudonymized ID numbers: Central Residence Register (age, sex, area of living, household size), Central Civil Status Register (civil status, death date, cause of death), and Educational Attainment Register (education). The study was approved by the Ethics Committee of the Medical University of Graz (EK-number: 1172/2024).

### Characteristics

We examined following socio-demographic measures: sex, education, area of living, household size, and civil status. Education was operationally defined as the highest degree obtained and differentiated between primary (International Standard Classification of Education; ISCED level 1-2), secondary (ISCED level 3-4) and tertiary education (ISCED level 5-8). We classified area of living according to the Degree of Urbanization Scheme [21] into cities, towns and suburbs, and rural areas.

Comorbidities listed as contributing causes on the death certificate were recorded in Austria only from 2019 onward using the International Classification of Diseases 10^th^ revision (ICD-10). These were categorized as: infectious and parasitic diseases (A00-B99), cancers (C00-C97), endocrine/nutritional/metabolic diseases (E00-E90), mental and behavioral disorders (F00-F99), diseases of the nervous system (G00-G99), diseases of the eye and adnexa and ear and mastoid process (H00-H95), circulatory diseases (I00-I99), respiratory diseases (J00-J99), digestive system diseases (K00-K93), diseases of the skin and subcutaneous tissue (L00-L99), musculoskeletal and connective tissue diseases (M00-M99), and genitourinary diseases (N00-N99) and analyzed when there were at least 10 observations present. Each comorbidity was coded as either present or absent. Given that mental and behavioral disorders constitute a major risk factor for suicide [8], we looked into specific diagnostic groups (i.e., organic mental disorders, affective disorders, and other ICD10 F-diagnoses) provided again that there were at least 10 observations present.

### Outcome

Suicide deaths were identified based on the ICD-10 codes (X60-X84, Y87.0). We differentiated between following suicide methods: poisoning (X60-69), hanging (X70), drowning (X71), firearms (X72-X74), jumping from a height (X80), and other methods (X75-79, X81-84, Y87.0).

### Statistical Analysis

First, sociodemographic characteristics (i.e., sex, education, civil status, household size, area of living) and comorbidities (i.e., mental and physical disorders) of older adults who died by suicide were compared with those who died by other causes than suicide. Differences were evaluated using chi-square tests and odds ratios (OR) together with their 95% confidence intervals (CI).

Second, we calculated the incidence rates of suicide based on the number of events and person-years for older adults in general as well as for the young-old (65-74 years), middle-old (75-84), and oldest-old (85 years and older). Further, we examined differences between the three age groups of older adults by using the young-old as the reference group and ORs, their 95% CI, chi-square trend tests, and linear by linear association tests. ORs were also calculated separately for each sex if there were at least 10 or more observations per combination. Lastly, we conducted a sensitivity analyses to reduce the effect of potential underreporting of suicides, where we also included deaths of undetermined intent (ICD-10: Y10-Y34, Y87.2) [22].

All analyses were performed using R 4.4.3 [23] and the R-packages *stats* (4.1.3) [23], *coin* (1.4.3) [24], and *MASS* (7.3.65) [25].

## Results

Between January 1, 2014 and December 31, 2023, a total of 724,072 individuals aged 65 and over died. Of these 4,724 (0.7%) died by suicide; most deaths were due to circulatory diseases (*n* = 302,939; 41.8%), cancers (*n* = 157,461; 21.7%), and respiratory diseases (*n* = 43,069; 5.9%). Compared to those who died by other causes, those who died by suicide were younger (suicide: 78.5 years, *SD* = 7.55; other causes: 83.7 years, *SD* = 8.41, *t*(4,800) = 47, *p* ≥ .001) and more likely to be male, higher educated, divorced, married, living alone, and living in towns and rural areas (see Supplementary Table 1). During 2019-2023, those who died by suicide were more likely to have a mental and behavioral disorder, especially a mood disorder, and eye and ear related diseases when compared to other decedents.

Among those who died by suicide, 1,680 (76.7% male) were young-old (65-74 years), 2,008 (77.7% male) were middle old (75-84 years), and 1,036 (77.9% male) were oldest-old (85 years and older). The overall suicide rate was 23.4 per 100,000 person-years. The highest rate was found among oldest-old (33.2; 95% CI = 29.5, 37.2), followed by middle-old (23.0; 95% CI = 21.8, 24.2) and the lowest rate was found among young-old adults (17.4; 95% CI = 16.8, 18.1). Across all age groups, suicide rates were significantly higher in males versus females (see Figure 1).

**Figure 1:**
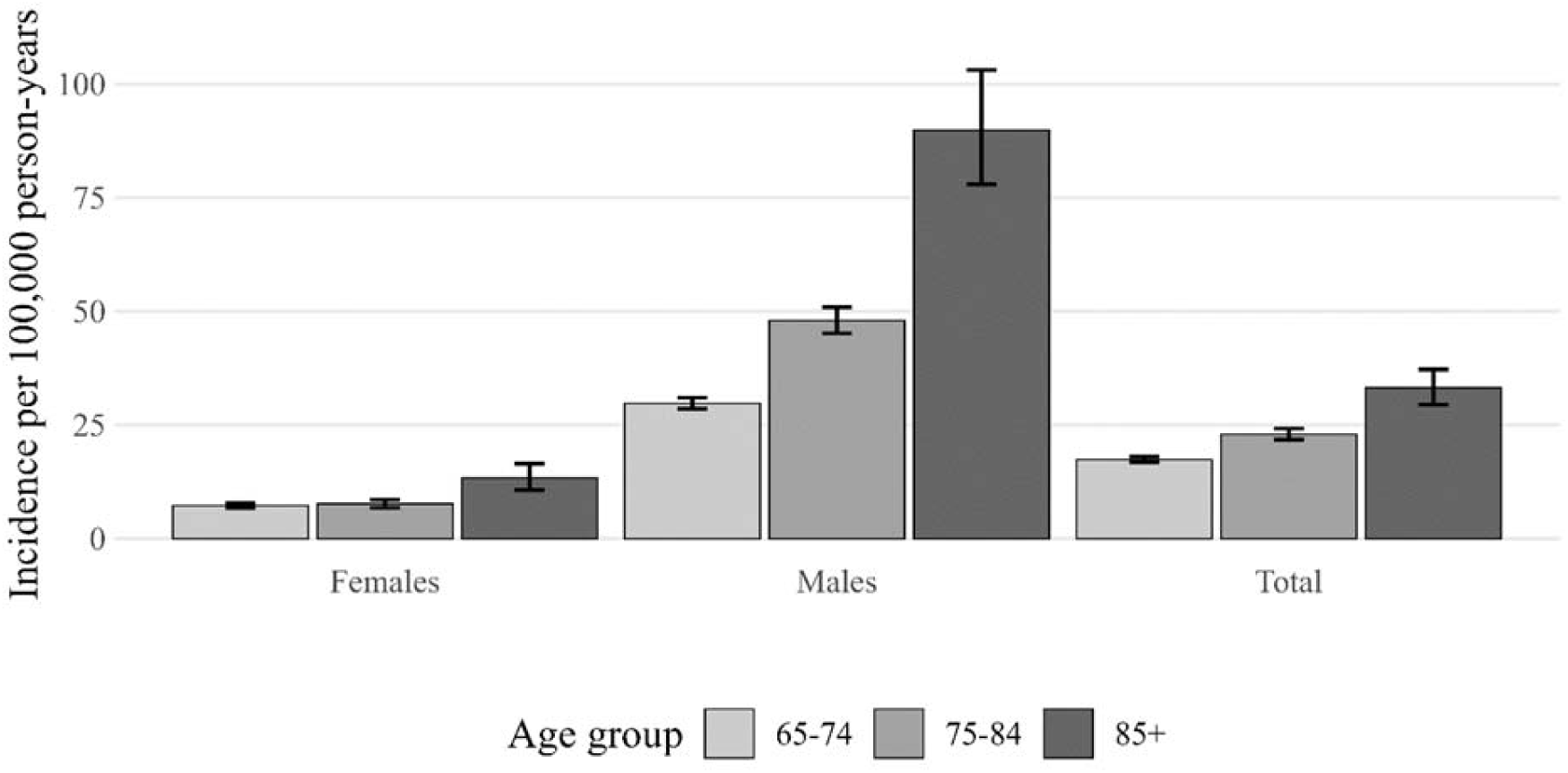
Crude incidence rate of suicide by sex and age group (per 100,000 person-years) together with 95% confidence intervals (assumed Poisson distribution of the event counts) between 2014-2023

The majority of young- and middle-old who died by suicide were married (53.5%, 50.5%, respectively), while the majority of oldest-old was widowed (51.8%; Table 1). Compared to the young-old, the middle-old were less likely to be single (OR = 0.6; 95% CI = 0.4, 0.7), less likely to be divorced (OR = 0.5; 95% CI = 0.4, 0.6), and more likely to be widowed (OR = 2.6; 95% CI =2.2, 3.1). Oldest-old were also less likely to be single (OR = 3.3; 95% CI = 0.2, 0.5), divorced (OR = 0.3; 95% CI = 0.2, 0.3) and married (OR = 0.5; 95% CI = 0.5,0.6), and more likely to be widowed (OR = 6.2; 95% CI = 5.2, 7.4) versus young-olds. Most suicides occurred by hanging (40.8%), followed by firearms (27.5%), jumping from a height (9.8%), poisoning (7.8%), and drowning (4.1%). While the proportion of those who died by hanging increased with age (χ^2^(1) = 8.0, *p* = .005), those by poisoning decreased (χ^2^(1) = 7.0, *p* = .01).

**Table 1:** Prevalence of different characteristics among young old, middle old, and oldest old who died by suicide. Overall age-group differences were assessed using trend-tests, unadjusted odds ratios (OR) and their 95% confidence intervals (95% CI).

| Full cohort (2012–2023) | Total<br>N = 4,724 | Young-old<br>n = 1,680 | Middle-old<br>n = 2,008 | Oldest-old<br>n = 1,036 | Trend test | OR (95% CI)<br>Young-old vs. Middle-old | OR (95% CI)<br>Young-old vs. Oldest-old |
| --- | --- | --- | --- | --- | --- | --- | --- |
| Sex |  |  |  |  |  |  |  |
| Female | 1,069 (22.6%) | 392 (23.3%) | 448 (22.3%) | 229 (22.1%) | $\chi^2 = 0.6, p = .400$ | 1.06 (0.91, 1.24) | 1.07 (0.89, 1.29) |
| Male | 3,655 (77.4%) | 1,288 (76.7%) | 1,560 (77.7%) | 807 (77.9%) |  |  |  |
| Education |  |  |  |  |  |  |  |
| Primary | 1,569 (33.2%) | 393 (23.4%) | 723 (36.0%) | 453 (43.7%) | $Z = -11, p \leq .001$ | 0.58 (0.51, 0.66) | 0.42 (0.36, 0.50) |
| Secondary | 2,864 (60.6%) | 1,160 (69.0%) | 1,172 (58.4%) | 532 (51.4%) |  |  |  |
| Tertiary | 291 (6.2%) | 127 (7.6%) | 113 (5.6%) | 51 (4.9%) |  |  |  |
| Marital Status |  |  |  |  |  |  |  |
| Divorced | 623 (13.2%) | 333 (19.8%) | 228 (11.4%) | 62 (6.0%) | $\chi^2 = 115, p \leq .001$ | 0.52 (0.43, 0.62) | 0.26 (0.19, 0.34) |
| Widowed | 1,414 (29.9%) | 249 (14.8%) | 628 (31.3%) | 537 (51.8%) | $\chi^2 = 419, p \leq .001$ | 2.62 (2.22, 3.09) | 6.18 (5.16, 7.43) |
| Married | 2,304 (48.8%) | 898 (53.5%) | 1,014 (50.5%) | 392 (37.8%) | $\chi^2 = 56, p \leq .001$ | 0.89 (0.78, 1.01) | 0.53 (0.45, 0.62) |
| Single | 383 (8.1%) | 200 (11.9%) | 138 (6.9%) | 45 (4.3%) | $\chi^2 = 54, p \leq .001$ | 0.55 (0.43, 0.68) | 0.34 (0.24, 0.46) |
| Area of living |  |  |  |  |  |  |  |
| Cities | 1,225 (25.9%) | 455 (27.1%) | 499 (24.9%) | 271 (26.2%) | $Z = 0.08, p = .900$ | 1.04 (0.92, 1.17) | 0.99 (0.86, 1.14) |
| Towns and suburbs | 1,473 (31.2%) | 499 (29.7%) | 645 (32.1%) | 329 (31.8%) |  |  |  |
| Rural | 2,026 (42.9%) | 726 (43.2%) | 864 (43.0%) | 436 (42.1%) |  |  |  |
| Living alone |  |  |  |  |  |  |  |
| No | 2,901 (61.4%) | 1,050 (62.5%) | 1,259 (62.7%) | 592 (57.1%) | $\chi^2 = 6, p = .010$ | 0.99 (0.987, 1.13) | 1.25 (1.07, 1.46) |
| Yes | 1,823 (38.6%) | 630 (37.5%) | 749 (37.2%) | 444 (42.9%) |  |  |  |
| Suicide methods |  |  |  |  |  |  |  |
| Drowning | 196 (4.1%) | 79 (4.7%) | 83 (4.1%) | 34 (3.3%) | $\chi^2 = 3, p = .070$ | 0.87 (0.64, 1.2) | 0.69 (0.45, 1.03) |
| Poisoning | 370 (7.8%) | 160 (9.5%) | 136 (6.8%) | 74 (7.1%) | $\chi^2 = 7, p = .010$ | 0.69 (0.54, 0.88) | 0.73 (0.55, 0.97) |
| Firearms | 1,298 (27.5%) | 427 (25.4%) | 602 (30.0%) | 269 (26.0%) | $\chi^2 = 0.7, p = .400$ | 1.26 (1.09, 1.45) | 1.03 (0.86, 1.23) |
| Hanging | 1,926 (40.8%) | 649 (38.6%) | 819 (40.8%) | 458 (44.2%) | $\chi^2 = 8, p = .005$ | 1.09 (0.96, 1.25) | 1.26 (1.08, 1.47) |
| Jumping from height | 465 (9.8%) | 172 (10.2%) | 179 (8.9%) | 114 (11.0%) | $\chi^2 = 0.1, p = .700$ | 0.86 (0.69, 1.07) | 1.08 (0.84, 1.39) |
| Other<sup>a</sup> | 469 (9.9%) | 193 (11.5%) | 189 (9.4%) | 87 (8.4%) | $\chi^2 = 8, p = .006$ | 0.80 (0.65, 0.99) | 0.71 (0.54, 0.92) |
| Restricted to 2019–2023 | Total<br>N = 2,392 | Young-old<br>n = 829 | Middle-old<br>n = 1,049 | Oldest-old<br>n = 514 | Trend test | OR (95% CI)<br>Young-old vs. Middle-old | OR (95% CI)<br>Young-old vs. Oldest-old |
| Cancers (C00-C97) | 305 (12.8%) | 105 (12.7%) | 147 (14.0%) | 53 (10.3%) | $\chi^2 = 1, p = 0.300$ | 1.12 (0.86, 1.47) | 0.79 (0.56, 1.12) |
| Endocrine/nutritional/metabolic (E00-E90) | 137 (5.7%) | 41 (4.9%) | 68 (6.5%) | 28 (5.4%) | $\chi^2 = 0.4, p = 0.500$ | 1.33 (0.90, 2.00) | 1.11 (0.67, 1.81) |
| Mental behavioral (F00-F99) | 581 (24.3%) | 222 (26.8%) | 253 (24.1%) | 106 (20.6%) | $\chi^2 = 7, p = .010$ | 0.87 (0.71, 1.07) | 0.71 (0.54, 0.92) |
| Organic mental disorders (F00-F09) | 56 (2.3%) | 12 (1.4%) | 24 (2.3%) | 20 (3.9%) | $\chi^2 = 8, p = .005$ | 1.59 (0.81, 3.32) | 2.76 (1.35, 5.85) |
| Affective disorders (F30-F39) | 498 (20.8%) | 192 (23.2%) | 218 (20.8%) | 88 (17.1%) | $\chi^2 = 7, p = .009$ | 0.87 (0.70, 1.08) | 0.69 (0.52, 0.90) |
| Other F diagnoses (F10-F29, F40-F99) | 98 (4.1%) | 48 (5.8%) | 36 (3.4%) | 14 (2.7%) | $\chi^2 = 9, p = .003$ | 0.58 (0.37, 0.90) | 0.46 (0.24, 0.81) |
| Nervous system (G00-G99) | 139 (5.8%) | 49 (5.9%) | 67 (6.4%) | 23 (4.5%) | $\chi^2 = 0.8, p = .400$ | 1.09 (0.74, 1.60) | 0.75 (0.44, 1.23) |
| Circulatory system (I00-I99) | 508 (21.2%) | 126 (15.2%) | 240 (22.9%) | 140 (27.2%) | $\chi^2 = 30, p \leq .001$ | 1.66 (1.31, 2.10) | 2.09 (1.59, 2.74) |
| Respiratory system (J00-J99) | 159 (6.6%) | 58 (7.0%) | 71 (6.8%) | 30 (5.8%) | $\chi^2 = 0.6, p = .400$ | 0.97 (0.67, 1.39) | 0.82 (0.52, 1.29) |
| Musculoskeletal system and connective tissue (M00-M99) | 73 (3.1%) | 14 (1.7%) | 38 (3.6%) | 21 (4.1%) | $\chi^2 = 7, p = .007$ | 2.19 (1.21, 4.20) | 2.48 (1.26, 5.03) |
| Genitourinary system (N00-N99) | 80 (3.3%) | 18 (2.2%) | 35 (3.3%) | 27 (5.3%) | $\chi^2 = 9, p = .003$ | 0.97 (0.67, 1.39) | 0.82 (0.52, 1.29) |

*Note.* Trend test refers either to a chi-squared test for trend ( $df = 1$ ) or a linear-by-linear associations test in case of ordered categories (i.e., education or area of living). Data on comorbid diagnoses were only available from 2019 onward; therefore, analyses involving comorbidities were restricted to 2019–2023.
<sup>a</sup> Other = ICD-10 codes X75-X79, X81-X84, and Y87.0

Mental and behavioral disorders were the most prevalent comorbid conditions among those who died by suicide (24.3%) (Table 1). Within this category, affective disorders (20.8%) were the most common. Relative with increasing age, we found that dementia and organic disorders became more prevalent among those who died by suicide (χ^2^(1) =8.0, *p* = .005). Likewise, circulatory diseases (χ^2^(1) = 30.0, *p* ≤ .001) and diseases of the genitourinary system (χ^2^(1) = 9.0, *p* = .003) also became more prevalent. Oldest-old who died by suicide were less likely to have a cancer diagnosis when compared to the young-olds (OR = 0.8; 95% CI = 0.6, 1.1), and more likely to have diagnoses related to musculoskeletal system and connective tissue (OR = 2.5; 95% CI = 1.3, 5.0).

Sex specific analyses (Table 2) found that the oldest-old individuals, versus young-old, were more likely to be widowed for both males (OR = 7.1; 95% CI = 5.7, 8.9) and females (OR = 6.5; 95% CI = 4.5, 9.3). Oldest-old males were also more likely to be living alone (OR = 1.24; 95% CI = 1.03, 1.49). Hanging was the most common suicide method for both males and females, followed by firearms and jumping from a height among males and by drug poisoning and jumping from a height among females. Compared to the young-olds, oldest-old males were more likely to die by hanging (OR = 1.2; 95% CI = 1.0, 1.5), while oldest-old females were more likely to die by jumping from a height (OR= 1.6; 95% CI = 1.0, 2.4). Circulatory diseases were more likely present in middle-old males (OR = 1.7; 95% CI = 1.3, 2.2), females (OR = 1.7; 95% CI = 1.0, 2.8) and oldest-old males (OR = 2.0; 95% CI = 1.5, 2.7) and females (OR = 2.6; 95% CI = 1.5, 4.6) compared to the young-olds. Oldest old males were less likely to have been diagnosed with a mental and behavioral disorder (OR = 0.71; 95% CI = 0.52, 0.96).

**Table 2:** Characteristics of suicide deaths stratified by age group and sex. Values are presented as numbers and column percentages. Overall age-group differences were assessed using trend tests, unadjusted odds ratios (OR) and their 95% confidence intervals (95% CI).

| Male | Total<br>N = 3,655 | Young-old<br>n = 1,288 | Middle-old<br>n = 1,560 | Oldest-old<br>n = 807 | Trend test | OR (95% CI)<br>Young-old vs. Middle-old | OR (95% CI)<br>Young-old vs. Oldest-old |
| --- | --- | --- | --- | --- | --- | --- | --- |
| Education |  |  |  |  |  |  |  |
| Primary | 1,054 (28.8) | 258 (20.0%) | 496 (31.8%) | 300 (37.2%) |  |  |  |
| Secondary | 2,380 (65.1) | 933 (72.4%) | 979 (62.8%) | 468 (58.0%) | Z = -9, p ≤ .001 | 0.57 (0.49, 0.67) | 0.46 (0.38, 0.55) |
| Tertiary | 221 (6.1) | 97 (7.5%) | 85 (5.4%) | 39 (4.8%) |  |  |  |
| Marital Status |  |  |  |  |  |  |  |
| Divorced | 469 (12.8%) | 247 (19.2%) | 177 (11.3%) | 45 (5.6%) | $\chi^2 = 87$, p ≤ .001 | 0.54 (0.44, 0.66) | 0.25 (0.18, 0.34) |
| Widowed | 909 (24.9%) | 139 (10.8%) | 397 (25.4%) | 373 (46.2%) | $\chi^2 = 329$, p ≤ .001 | 2.82 (2.29, 3.49) | 7.10 (5.69, 8.91) |
| Married | 1996 (54.6%) | 747 (58.0%) | 890 (57.1%) | 359 (44.5%) | $\chi^2 = 31$, p ≤ .001 | 0.96 (0.83, 1.12) | 0.58 (0.49, 0.69) |
| Single | 2810 (7.7%) | 155 (12.0%) | 96 (6.2%) | 30 (3.7%) | $\chi^2 = 54$, p ≤ .001 | 0.48 (0.37, 0.62) | 0.28 (0.19, 0.42) |
| Area of living |  |  |  |  |  |  |  |
| Cities | 855 (23.4%) | 315 (24.5%) | 349 (22.4%) | 191 (23.7%) |  |  |  |
| Towns and suburbs | 1,170 (32.0%) | 382 (29.7%) | 524 (33.6%) | 264 (32.7%) | Z = -0.4, p = .700 | 0.99 (0.86, 1.14) | 0.96 (0.81, 1.13) |
| Rural | 1,630 (44.6%) | 591 (45.9%) | 687 (44.0%) | 352 (43.6%) |  |  |  |
| Living alone |  |  |  |  |  |  |  |
| No | 2,406 (65.8%) | 854 (66.3%) | 1,057 (67.8%) | 495 (61.3%) |  |  |  |
| Yes | 1,249 (34.2%) | 434 (33.7%) | 503 (32.2%) | 312 (38.7%) | $\chi^2 = 4$, p = .050 | 0.94 (0.8, 1.1) | 1.24 (1.03, 1.49) |
| Suicide methods |  |  |  |  |  |  |  |
| Drowning | 86 (2.4%) | 29 (2.3%) | 42 (2.7%) | 15 (1.9%) | $\chi^2 = 0.2$, p = .700 | 1.2 (0.75, 1.96) | 0.82 (0.43, 1.52) |
| Poisoning | 149 (4.1%) | 69 (5.4%) | 48 (3.1%) | 32 (4.0%) | $\chi^2 = 4$, p = .050 | 0.56 (0.38, 0.81) | 0.73 (0.47, 1.11) |
| Firearms | 1,258 (34.4%) | 403 (31.3%) | 589 (37.8%) | 266 (33.0%) | $\chi^2 = 2$, p = .200 | 1.33 (1.14, 1.56) | 1.08 (0.89, 1.30) |
| Hanging | 1,544 (42.2%) | 525 (40.8%) | 650 (41.7%) | 369 (45.7%) | $\chi^2 = 4$, p = .030 | 1.04 (0.89, 1.21) | 1.22 (1.03, 1.46) |
| Jumping from height | 290 (7.9%) | 119 (9.2%) | 102 (6.5%) | 69 (8.6%) | $\chi^2 = 0.9$, p = .300 | 0.69 (0.52, 0.90) | 0.92 (0.67, 1.25) |
| Other<sup>a</sup> | 328 (9.0%) | 143 (11.1%) | 129 (8.3%) | 56 (6.9%) | $\chi^2 = 12$, p ≤ .001 | 0.72 (0.56, 0.93) | 0.60 (0.43, 0.82) |
| Comorbidities<sup>b</sup> |  |  |  |  |  |  |  |
| Cancers (C00-C97) | 246 (13.2%) | 82 (13.0%) | 121 (14.9%) | 43 (10.3%) | $\chi^2 = 0.9$, p = .300 | 1.17 (0.87, 1.59) | 0.77 (0.52, 1.14) |
| Endocrine/nutritional/metabolic (E00-E90) | 114 (6.1%) | 34 (5.4%) | 58 (7.1%) | 22 (5.3%) | $\chi^2 = 0.03$, p = .900 | 1.35 (0.88, 2.11) | 0.98 (0.56, 1.69) |
| Mental behavioral (F00-F99) | 407 (21.9%) | 152 (24.1%) | 179 (22.0%) | 76 (18.3%) | $\chi^2 = 5$, p = .030 | 0.89 (0.70, 1.14) | 0.71 (0.52, 0.96) |
| Affective disorders (F30-F39) | 335 (18.0%) | 127 (20.1%) | 146 (18.0%) | 62 (14.9%) | $\chi^2 = 5$, p = .030 | 0.87 (0.67, 1.13) | 0.70 (0.50, 0.97) |
| Other F diagnoses (F00-F29, F40-F99) | 108 (3.5 %) | 37 (5.9%) | 46 (5.7%) | 25 (6.0%) | $\chi^2 = 0.005$, p = .900 | 0.96 (0.62, 1.51) | 1.03 (0.60, 1.73) |
| Nervous system (G00-G99) | 89 (4.8%) | 31 (4.9%) | 41 (5.0%) | 17 (4.1%) | $\chi^2 = 0.3$, p = .600 | 1.03 (0.64, 1.67) | 0.83 (0.44, 1.49) |
| Circulatory system (I00-I99) | 379 (20.4%) | 93 (14.7%) | 180 (22.1%) | 106 (25.5%) | $\chi^2 = 20$, p ≤ .001 | 1.65 (1.25, 2.18) | 1.98 (1.45, 2.71) |
| Respiratory system (J00-J99) | 118 (6.3%) | 42 (6.6%) | 52 (6.4%) | 24 (5.8%) | $\chi^2 = 0.3$, p = .600 | 0.96 (0.63, 1.47) | 0.86 (0.51, 1.43) |
| Genitourinary system (N00-N99) | 62 (3.3%) | 13 (2.1%) | 29 (3.6%) | 20 (4.8%) | $\chi^2 = 6$, p = .010 | 0.96 (0.63, 1.47) | 0.86 (0.51, 1.43) |
| Female | Total<br>N = 1,069 | Young-old<br>n = 392 | Middle-old<br>n = 448 | Oldest-old<br>n = 229 | Trend test | OR (95% CI)<br>Young-old vs. Middle-old | OR (95% CI)<br>Young-old vs. Oldest-old |
| Education |  |  |  |  |  |  |  |
| Primary | 515 (48.2%) | 135 (34.4%) | 227 (50.7%) | 153 (66.8%) |  |  |  |
| Secondary | 484 (45.3%) | 227 (57.9%) | 193 (43.1%) | 64 (27.9%) | Z = -7, p ≤ .001 | 0.55 (0.42, 0.72) | 0.29 (0.2, 0.40) |
| Tertiary | 70 (6.5%) | 30 (7.7%) | 28 (6.2%) | 12 (5.2%) |  |  |  |
| Marital Status |  |  |  |  |  |  |  |
| Divorced | 154 (14.4%) | 86 (21.9%) | 51 (11.4%) | 17 (7.4%) | $\chi^2 = 28$, p ≤ .001 | 0.46 (0.31, 0.66) | 0.29 (0.16, 0.48) |
| Widowed | 505 (47.2%) | 110 (28.1%) | 231 (51.6%) | 164 (71.6%) | $\chi^2 = 116$, p ≤ .001 | 2.73 (2.05, 3.65) | 6.47 (4.53, 9.34) |
| Married | 308 (28.8%) | 151 (38.5%) | 124 (27.7%) | 33 (14.4%) | $\chi^2 = 41, p \leq .001$ | 0.61 (0.46, 0.82) | 0.27 (0.17, 0.40) |
| Single | 102 (9.5%) | 45 (11.5%) | 42 (9.4%) | 15 (6.6%) | $\chi^2 = 4, p = .040$ | 0.80 (0.51, 1.24) | 0.54 (0.29, 0.97) |
| Area of living |  |  |  |  |  |  |  |
| Cities | 370 (34.6%) | 140 (35.7%) | 150 (33.5%) | 80 (34.9%) |  |  |  |
| Towns and suburbs | 303 (28.3%) | 117 (29.8%) | 121 (27.0%) | 65 (28.4%) | $Z = 0.6, p = 0.500$ | 1.17 (0.91, 1.51) | 1.07 (0.79, 1.44) |
| Rural | 396 (37.0%) | 135 (34.4%) | 177 (39.5%) | 84 (36.7%) |  |  |  |
| Living alone |  |  |  |  |  |  |  |
| No | 495 (46.3%) | 196 (50.0%) | 202 (45.1%) | 97 (42.4%) |  |  |  |
| Yes | 874 (53.7%) | 196 (50.0%) | 246 (54.9%) | 132 (57.6%) | $\chi^2 = 4, p = .050$ | 1.22 (0.93, 1.60) | 1.36 (0.98, 1.89) |
| Suicide methods |  |  |  |  |  |  |  |
| Drowning | 110 (10.3%) | 50 (12.8%) | 41 (9.2%) | 19 (8.3%) | $\chi^2 = 4, p = .060$ | 0.69 (0.44, 1.07) | 0.62 (0.35, 1.06) |
| Poisoning | 221 (20.7%) | 91 (23.2%) | 88 (19.6%) | 42 (18.3%) | $\chi^2 = 2, p = .100$ | 0.81 (0.58, 1.13) | 0.74 (0.49, 1.11) |
| Hanging | 382 (35.7%) | 124 (31.6%) | 169 (37.7%) | 89 (38.9%) | $\chi^2 = 4, p = .050$ | 1.31 (0.98, 1.74) | 1.37 (0.98, 1.93) |
| Jumping from height | 175 (16.4%) | 53 (13.5%) | 77 (17.2%) | 45 (19.7%) | $\chi^2 = 4, p = .040$ | 1.33 (0.91, 1.95) | 1.56 (1.01, 2.42) |
| Other<sup>c</sup> | 181 (16.9%) | 74 (18.9%) | 73 (16.3%) | 34 (14.8%) | $\chi^2 = 2, p = .200$ | 0.84 (0.59, 1.19) | 0.75 (0.48, 1.16) |
| Comorbidities<sup>d</sup> |  |  |  |  |  |  |  |
| Cancers (C00-C97) | 59 (11.1%) | 23 (11.7%) | 26 (11.0%) | 10 (10.2%) | $\chi^2 = 0.1, p = .700$ | 0.94 (0.52, 1.71) | 0.86 (0.38, 1.84) |
| Mental behavioral (F00-F99) | 174 (32.8%) | 70 (35.5%) | 74 (31.4%) | 30 (30.6%) | $\chi^2 = 0.9, p = .300$ | 0.83 (0.55, 1.24) | 0.80 (0.47, 1.34) |
| Affective disorders (F30-F39) | 163 (30.7) | 65 (33.3%) | 72 (30.5%) | 26 (26.5%) | $\chi^2 = 1, p = .300$ | 0.89 (0.59, 1.34) | 0.73 (0.42, 1.25) |
| Circulatory system (I00-I99) | 127 (23.9%) | 33 (16.8%) | 60 (25.4%) | 34 (34.7%) | $\chi^2 = 12, p \leq .001$ | 1.69 (1.06, 2.75) | 2.64 (1.51, 4.64) |

*Note.* Trend test refers either to a chi-squared test for trend ( $df = 1$ ) or a linear-by-linear associations test in case of ordered categories (i.e., education or area of living).
<sup>a</sup> Other = ICD-10 codes X75-X79, X81-X86, and Y87.0
<sup>b</sup> Data on comorbid diagnoses were only available from 2019 onward; therefore, analyses involving comorbidities were restricted to 2019–2023; $n_{\text{total}} = 1,861$ , $n_{\text{young-old}} = 632$ , $n_{\text{middle-old}} = 813$ , $n_{\text{oldest-old}} = 416$
<sup>c</sup> Other = ICD-10 codes X72-X79, X81-X84, and Y87.0
<sup>d</sup> Data on comorbid diagnoses were only available from 2019 onward; therefore, analyses involving comorbidities were restricted to 2019–2023; $n_{\text{total}} = 531$ , $n_{\text{young-old}} = 197$ , $n_{\text{middle-old}} = 263$ , $n_{\text{oldest-old}} = 98$

In a sensitivity analysis (Supplementary Table 2), we included those of undetermined intent (1,005 cases) to those who died by suicide. Deaths of undetermined intent represented respectively 17.9%, 17.6%, and 16.8% of suicides across the three age groups and the overall pattern of associations remained largely unchanged.

## Discussion

In this nationwide, retrospective cohort study we found differences in suicide rates of older adults, with the oldest-old having the highest suicide rate. We further found that low education, widowhood, diseases related to musculoskeletal system and connective tissue, and circulatory diseases were more common among middle-old and oldest-old compared with young-old suicide decedents.

The high suicide rates among the oldest-old, especially males, is supported by the WHO Mortality Database [10]. International evidence regarding the potential protective role of marriage on risks of suicide in older adults is mixed [19,26,27]. We also found that oldest-old who died by suicide were less likely to be married. Older adults who live as singles may have smaller social networks, as suggested for those never married [28]. While network size does not directly equate to the quality of social support, smaller networks might indicate limited access to supportive relationships. This may be particularly consequential for males, who are less likely to receive emotional support through conversations compared to females or to expand their networks after the loss of a partner [29,30]. Consequently, males may experience greater loneliness, lower well-being, and poorer adjustment to bereavement, which may impair adaptive coping with additional stressors such as declining health, ultimately contributing to suicide [30–34]. Widowhood is more common among the oldest-old and seems a relevant risk factor for suicide in later life [17]. Again, it should be noted that the higher odds of widowhood found in this study may, at least in part, reflect the higher prevalence of widowhood in old age.

Firearms is one of the most lethal methods of suicide [35] and the second most frequently used method among older adults in Austria. We found that the proportion who died by hanging, generally considered a ‘determined’ method [36], increased relative with increasing age [10]. In contrast, the proportion of less determined methods [36] such as poisoning, which carry a higher chance of rescue, intervention and survival, declined with age. It is important to note, however, that suicide by poisoning may go undetected. Poisoning deaths may be misclassified as deaths of undetermined intent, accidents, or drug use disorder deaths [37] rather than suicides, particularly when toxicological (e.g., autopsy rate was below European average^1^ with 5.8% for all deaths and 8.9% for suicides) or other evidence of suicidal intent (e.g., suicide notes) is lacking [37–39].

The increasing prevalence of cardiovascular and musculoskeletal system and connective tissue diseases with increasing age partly align with Australian evidence [18]. Cancer being less common among the oldest-old than younger age groups is also supported by other findings [40]. The higher (e.g., for a cardiovascular disease) or lower odds (e.g., for a cancer diagnosis) likely reflect the higher or lower age-related prevalence of these health conditions.

### Implications

As partner loss either through divorce or widowhood is often regarded as one of the most profound forms of relational loss [41], interventions that support partners after the loss in coping with this transition could play a role in reducing suicide risk. For example, the community-based intervention by Oyama and colleagues [42], while not specifically targeting widowed individuals, demonstrated a significant reduction in suicide rates among female older adults through social group activities and psychoeducational support. Likewise, Davidow and colleagues [43] identified multiple effective group and individual interventions specifically designed for older adults experiencing spousal bereavement, demonstrating that such approaches can effectively address grief-related distress and mental health outcomes.

Despite widowhood, further psychosocial factors, such as, social isolation, and loneliness [46], may be equally important contributors to suicide risk, as the accumulation of social and emotional stressors can weaken coping capacities and diminish perceived belonging, which in turn can increase suicide risk among older adults [8]. Accordingly, suicide prevention programs in later life should extend beyond the treatment of affective disorders to include community-based programs that foster social engagement and support [7,47]. Many available interventions have mainly focused on depression and isolation, and their effectiveness has been found to differ by gender, with better outcomes observed among women [47]. Therefore, prevention strategies for older men, a most vulnerable group, may require alternative approaches or settings. Because suicide cases were more often residing in non-urban areas (i.e., towns and suburbs, and rural areas), similar to a previous study [48], any strategies to prevent suicides at older age should be considered for implementation in rural areas to reduce suicide rates among older adults.

A key and effective suicide prevention strategy is restricting access to lethal means. The implementation of restriction measures, ranging from limiting firearm access to securing buildings and bridges, was found to decrease suicides [9]. The firearm legislation reform in particular, which imposed restrictions for instance, on the purchase, access, or use of firearms, was shown to influence suicide deaths by firearm in certain countries [44]. In Austria, the tightening of the firearm legislation in 1997 was associated with a decline in firearm suicides, particularly among males older than 65 years [45]. Hence, further tightening of firearm access could be useful.

### Strengths and limitations

Strengths of this study include using national individual-level data on all suicide deaths. Limitations include, that in Austria, physicians can optionally report additional significant health conditions on the medical death certificates, i.e., conditions that contributed to death but were not directly related to the underlying cause, but they are not required to do so. This likely results in underreporting of, for instance, affective disorders [49], therefore limiting the precision of analyses linking comorbidities to suicide deaths [50].

## Conclusion

In this national, retrospective cohort study, we find distinct differences in suicide mortality between young-old, middle-old, and oldest-old older adults in Austria. The oldest-old tend to use more lethal methods compared to young-old. Middle-old individuals were less likely to be divorced, while oldest-old individuals were more likely to be widowed, reflecting not only the increased likelihood of spousal loss with advancing age but also highlighting an area where intervention could be beneficial. Targeted measures, such as bereavement support for the oldest-old, might help reduce suicide risks among older adults.

## Acknowledgements

This research project was conducted with data from the Austrian Micro Data Center (AMDC). The AMDC is a research data infrastructure facility of Statistics Austria that enables research on micro data processed in compliance with data protection regulations.

## Data

Access and linkage of register data was approved of and provided by the Austrian Micro Data Center (AMDC) of Statistics Austria following national legislation (Bundesstatistikgesetz §31, Forschungsorganisationsgesetz §38b). The AMDC is a research data infrastructure facility of Statistics Austria that enables research on micro data processed in compliance with data protection regulations. The data used for this research can be accessed by researchers at scientific institutions accredited with the AMDC against a fee. For further information, see https://www.statistik.at/en/services/tools/services/center-for-science/austrian-micro-data-center-amdc

## Abbreviations

AMDC: Austria Micro Data Center
ISCED: International Standard Classification of Education
ICD-10: International Classification of Diseases and Related Health Problems
OR: Odds Ratio
95% CI: 95% confidence interval

## Footnotes

1 https://gateway.euro.who.int/en/hfa-explorer/

