## Supplement for "Suicide among older adults in Austria"

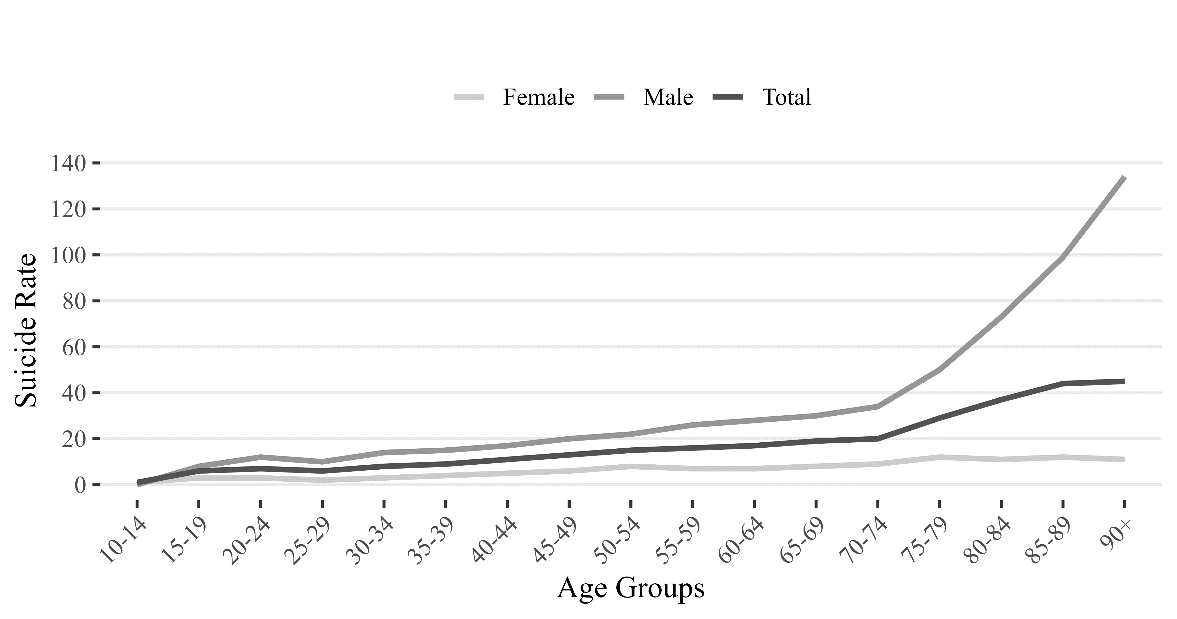
*Supplementary Figure 1: Standardized suicide rate per 100.000 persons per age-group (five-year average 2019-2023)*

*Note*. Standardized suicide rates per 100,000 persons (using the 2013 European Standard Population). Figure adapted from *Suizid und Suizidprävention Österreich. Bericht 2025* by Bundesministerium für, Soziales, Gesundheit, Pflege und Konsumentenschutz (BMSGPK) (Ed.), 2025.[51] Data was obtained from GÖG (<https://goeg.at/>).

*Supplementary Table 1: Demographic and health-related characteristics by outcome (death by suicide, death by other cause). Values are presented as numbers and column percent. Overall group differences were assessed using chi-square tests. Unadjusted odds ratios (OR) and their 95% confidence intervals (95% CI) compare other death vs. suicide.*

| **Full cohort (2012–2023)** | **Total**  ***N* = 724,072** | **Other death**  ***n* = 719,348** | **Suicide**  ***n* = 4,724** | **χ^2^ (*df*), p value** | **OR (95% CI) Other death vs. Suicide** |
| --- | --- | --- | --- | --- | --- |
| Sex |  |  |  |  |  |
| Female | 390,806 (54.0%) | 389,737 (54.2%) | 1,069 (22.6%) | 1,879 (1), p ≤ .001 | 4.04 (3.78, 4.33) |
| Male | 333,266 (46.0%) | 329,611 (45.8%) | 3,655 (77.4%) |  |  |
| Education |  |  |  |  |  |
| Primary | 333,130 (46.0%) | 331,561 (46.1%) | 1,569 (33.2%) | 313 (2), p ≤ .001 | 1.63 (1.54, 1.72) |
| Secondary | 354,622 (49.0%) | 351,758 (48.9%) | 2,864 (60.6%) |  |  |
| Tertiary | 36,320 (5.0%) | 36,029 (5.0%) | 291 (6.2%) |  |  |
| Marital Status |  |  |  |  |  |
| Divorced | 64,923 (9.0%) | 64,300 (8.9%) | 623 (13.2%) | 103 (1), p ≤ .001 | 1.55 (1.42, 1.68) |
| Widowed | 324,701 (44.8%) | 323,287 (44.9%) | 1,414 (29.9%) | 427 (1), p ≤ .001 | 0.52 (0.49, 0.56) |
| Married | 277,767 (38.4%) | 275,463 (38.3%) | 2,304 (48.8%) | 217 (1), p ≤ .001 | 1.53 (1.45, 1.62) |
| Single | 56,681 (7.8%) | 56,298 (7.8%) | 383 (8.1%) | 0.5 (1), p = .500 | 1.04 (0.93, 1.15) |
| Area of living |  |  |  |  |  |
| Cities | 226,071 (31.2%) | 224,846 (31.3%) | 1,225 (25.9%) | 88 (2), p ≤ .001 | 1.29 (1.22, 1.36) |
| Towns and suburbs | 230,210 (31.8%) | 228,737 (31.8%) | 1,473 (31.2%) |  |  |
| Rural | 267,791 (37.0%) | 265,765 (36.9%) | 2,026 (42.9%) |  |  |
| Living alone |  |  |  |  |  |
| No | 489,396 (67.6%) | 486,495 (67.6%) | 2,901 (61.4%) | 83 (1), p ≤ .001 | 1.31 (1.24, 1.39) |
| Yes | 234,676 (32.4%) | 232,853 (32.4%) | 1,823 (38.6%) |  |  |
| **Restricted to 2019–2023** | **Total**  ***N* = 381,990** | **Other death**  ***n* = 379,598** | **Suicide**  ***n* = 2,392** | **χ^2^(*df*), p value** | **OR (95% CI) Other death vs. Suicide** |
| Infectious and parasitic diseases (A00-B99) | 37,844 (9.9%) | 37,821 (10.0%) | 23 (1.0%) | 215 (1), p ≤ .001 | 0.09 (0.06, 0.13) |
| Cancers (C00-C97) | 96,202 (52.2%) | 95,897 (25.3%) | 305 (12.8%) | 179 (1), p ≤ .001 | 0.43 (0.38, 0.49) |
| Endocrine/ nutritional/metabolic (E00-E90) | 83,429 (21.8%) | 83,292 (21.9%) | 137 (5.7%) | 365 (1), p ≤ .001 | 0.22 (0.18, 0.26) |
| Mental behavioral (F00-F99) | 67,389 (17.6%) | 66,808 (17.6%) | 581 (24.3%) | 73 (1), p ≤ .001 | 1.50 (1.37, 1.65) |
| F00-F09 | 55,448 (14.5%) | 55,392 (14.6%) | 56 (2.3%) | 287 (1), p ≤ .001 | 0.14 (0.11, 0.18) |
| F10-F19 | 7,438 (1.9%) | 7,395 (1.9%) | 43 (1.8%) | 0.2 (1), p =.600 | 0.92 (0.67, 1.23) |
| F20-F29 | 1,370 (0.4%) | 1,351 (0.4%) | 19 (0.8%) | 12 (1), p ≤ .001 | 2.24 (1.37, 3.43) |
| F30-F39 | 5,447 (1.4%) | 4,949 (1.3%) | 498 (20.8%) | 6,427 (1), p ≤ .001 | 19.90 (17.95, 22.04) |
| F40-F48 | 505 (0.1%) | 447 (0.1%) | 28 (1.2%) | 189 (1), p ≤ .001 | 9.41 (6.27, 13.54) |
| F50-F99 | 790 (0.2%) | 779 (0.2%) | 11 (0.5%) | 6 (1), p = .010 | 2.25 (1.16, 3.87) |
| Nervous system (G00-G99) | 48,154 (12.6%) | 48,015 (12.6%) | 139 (5.8%) | 100 (1), p ≤ .001 | 0.43 (0.36, 0.50) |
| Eye/ear (H00-H95) | 1,628 (0.4%) | 1,607 (0.4%) | 21 (0.9%) | 11 (1), p = .001 | 2.08 (1.31, 3.12) |
| Circulatory system (I00-I99) | 276,895 (72.5%) | 276,389 (72.8%) | 506 (21.2%) | 3,178 (1), p ≤ .001 | 0.10 (0.09, 0.11) |
| Respiratory system (J00-J99) | 118,303 (31.0%) | 118,144 (31.1%) | 159 (6.6%) | 665 (1), p ≤ .001 | 0.16 (0.13, 0.18) |
| Digestive system (K00-K93) | 37,811 (9.9%) | 37,751 (9.9%) | 60 (2.5%) | 147 (1), p ≤ .001 | 0.23 (0.18, 0.30) |
| Skin (L00-L99) | 7,269 (1.9%) | 7,253 (1.9%) | 16 (0.7%) | 19 (1), p ≤ .001 | 0.35 (0.20, 0.55) |
| Musculoskeletal system (M00-M99) | 18,017 (4.7%) | 17,944 (4.7%) | 73 (3.1%) | 14 (1), p ≤ .001 | 0.63 (0.50, 0.79) |
| Genitourinary system (N00-N99) | 85,637 (22.4%) | 85,557 (22.5%) | 80 (3.3%) | 502 (1), p ≤ .001 | 0.16 (0.13, 0.18) |

*Note*. Data on comorbid diagnoses were only available from 2019 onward; therefore, analyses involving comorbidities were restricted to 2019–2023.

*Supplementary Table 2: Demographic and health-related characteristics of individuals who died by suicide or undetermined intent (Y10-Y34, Y87.2), stratified by age-group. Values are presented as numbers and column percentages. Comparisons between age groups include trend tests, unadjusted odds ratios (OR) and their 95% confidence intervals (95% CI).*

| **Full cohort (2012–2023)** | **Total**  ***N* = 5,729** | **Young-old**  ***n* = 2,047** | **Middle-old**  ***n* = 2,437** | **Oldest-old**  ***n* = 1,245** | **Trend test** | **OR (95% CI) Young-old vs. Middle-old** | **OR (95% CI) Young-old vs. Oldest-old** |
| --- | --- | --- | --- | --- | --- | --- | --- |
| Sex |  |  |  |  |  |  |  |
| Female | 1,472 (25.7%) | 530 (25.9%) | 613 (25.2%) | 329 (26.4%) | χ^2^ = 0.04, p = .800 | 1.04 (0.91, 1.19) | 0.97 (0.83, 1.14) |
| Male | 4,257 (74.3%) | 1,517 (74.1%) | 1,824 (74.8%) | 916 (73.6%) |  |  |  |
| Education |  |  |  |  |  |  |  |
| Primary | 1,910 (33.3) | 477 (23.3%) | 888 (36.4%) | 545 (43.8%) | Z = -12, p ≤ .001 | 0.56 (0.49, 0.63) | 0.42 (0.36, 0.48) |
| Secondary | 3,444 (60.1%) | 1,397 (68.2%) | 1,409 (57.8%) | 638 (51.2%) |  |  |  |
| Tertiary | 375 (6.6%) | 173 (8.5%) | 140 (5.7%) | 62 (5.0%) |  |  |  |
| Marital Status |  |  |  |  |  |  |  |
| Divorced | 791 (13.8%) | 430 (21.0%) | 288 (11.8%) | 73 (5.9%) | χ^2^ = 160, p ≤ .001 | 0.50 (0.43, 0.59) | 0.23 (0.18, 0.30) |
| Widowed | 1,751 (30.6%) | 320 (15.6%) | 772 (31.7%) | 659 (52.9%) | χ^2^ = 506, p ≤ .001 | 2.50 (2.16, 2.90) | 6.07 (5.16, 7.15) |
| Married | 2,674 (46.7%) | 1,027 (50.2%) | 1,195 (49.0%) | 452 (36.3%) | χ^2^ = 51, p ≤ .001 | 0.96 (0.85, 1.07) | 0.57 (0.49, 0.65) |
| Single | 513 (9%) | 270 (13.2%) | 182 (7.5%) | 61 (4.9%) | χ^2^ = 73, p ≤ .001 | 0.53 (0.44, 0.65) | 0.34 (0.25, 0.45) |
| Area of living |  |  |  |  |  |  |  |
| Cities | 1,638 (28.6) | 607 (29.7%) | 679 (27.9%) | 352 (28.3%) | Z = 0.3, p = .800 | 1.17 (0.91, 1.51) | 1.07 (0.79, 1.44) |
| Towns and suburbs | 1,767 (30.8) | 601 (29.4%) | 775 (31.8%) | 391 (31.4%) |  |  |  |
| Rural | 2,324 (40.6) | 839 (41.0%) | 983 (40.3%) | 502 (40.3%) |  |  |  |
| Living alone |  |  |  |  |  |  |  |
| No | 3,422 (59.7%) | 1,240 (60.6%) | 1,490 (61.1%) | 692 (55.6%) | χ^2^ = 6, p = .010 | 0.98 (0.87, 1.10) | 1.23 (1.06, 1.42) |
| Yes | 2,307 (40.3%) | 807 (39.4%) | 947 (38.9%) | 553 (44.4%) |  |  |  |
| **Restricted to 2019–2023** | **Total**  ***N* = 3,002** | **Young-old**  ***n* = 1,037** | **Middle-old**  ***n* = 1,313** | **Oldest-old**  ***n* = 652** | **Trend test** | **OR (95% CI) Young-old vs. Middle-old** | **OR (95% CI) Young-old vs. Oldest-old** |
| Cancers (C00-C97) | 340 (11.3%) | 117 (11.3%) | 166 (12.6%) | 57 (8.7%) | χ^2^ = 2, p = .200 | 1.14 (0.89, 1.47) | 0.75 (0.54, 1.05) |
| Endocrine/nutritional/metabolic (E00-E90) | 192 (6.4%) | 60 (5.8%) | 92 (7.0%) | 40 (6.1%) | χ^2^ = 0.2, p = .600 | 1.23 (0.88, 1.72) | 1.06 (0.70, 1.60) |
| Mental behavioral (F00-F99) | 661 (22.0%) | 258 (24.9%) | 279 (21.2%) | 124 (19.0%) | χ^2^ = 9, p = .003 | 0.81 (0.67, 0.99) | 0.71 (0.56, 0.90) |
| Organic mental disorders (F00-F09) | 89 (3.0%) | 21 (2.0%) | 35 (2.7%) | 33 (5.1%) | χ^2^ = 12, p ≤ .001 | 1.32 (0.77, 2.33) | 2.58 (1.49, 4.56) |
| Affective disorders (F30-F39) | 529 (17.6%) | 209 (20.2%) | 227 (17.3%) | 93 (14.3%) | χ^2^ = 10, p = .002 | 0.83 (0.67, 1.02) | 0.66 (0.50, 0.86) |
| Other F diagnoses (F10-F29, F40-F99) | 123 (4.1%) | 63 (6.1%) | 45 (3.4%) | 15 (2.3%) | χ^2^ = 16, p ≤ .001 | 0.55 (0.37, 0.81) | 0.36 (0.20, 0.63) |
| Nervous system (G00-G99) | 181 (6.0%) | 65 (6.3%) | 89 (6.8%) | 27 (4.1%) | χ^2^ = 2, p = .100 | 1.09 (0.78, 1.52) | 0.65 (0.40, 1.01) |
| Circulatory system (I00-I99) | 731 (24.4%) | 183 (17.6%) | 349 (26.6%) | 199 (30.5%) | χ^2^ = 40, p ≤ .001 | 1.69 (1.38, 2.07) | 2.05 (1.63, 2.58) |
| Respiratory system (J00-J99) | 251 (8.4%) | 90 (8.7%) | 108 (8.2%) | 53 (8.1%) | χ^2^ = 0.2, p = .700 | 0.94 (0.70, 1.27) | 0.93 (0.65, 1.32) |
| Musculoskeletal system and connective tissue (M00-M99) | 84 (2.8%) | 15 (1.4%) | 45 (3.4%) | 24 (3.7%) | χ^2^ = 9, p = .003 | 2.42 (1.37, 4.51) | 2.60 (1.37, 5.11) |
| Genitourinary system (N00-N99) | 118 (3.9%) | 23 (2.2%) | 52 (4.0%) | 43 (6.6%) | χ^2^ = 20, p ≤ .001 | 0.94 (0.70, 1.27) | 0.93 (0.65, 1.32) |

*Note*. Trend test refers either to a chi-squared test for trend (*df* = 1) or a linear-by-linear associations test in case of ordered categories (i.e., education or area of living).

Data on comorbid diagnoses were only available from 2019 onward; therefore, analyses involving comorbidities were restricted to 2019–2023.
